# Knowledge and experiences of adolescent girls and young women in the use of sexual reproductive health and HIV services at health facilities in Maputo City, Mozambique

**DOI:** 10.1101/2024.09.14.24313688

**Authors:** Vasco A. Muchanga, Luisa Huo, Kathryn T. Kampa, Baltazar Chilundo, Khátia R. Munguambe, Troy D. Moon

## Abstract

**Background:** Knowledge and use of sexual reproductive health and human immunodeficiency virus (SRH and HIV) services are crucial for the prevention of pregnancy and sexually transmitted infections (STIs) among adolescent girls and young women (AGYW). This study aims to assess the knowledge and perceptions of AGYW about the SRH and HIV services offered in health facilities in Maputo, Mozambique.

**Material and methods:** A cross-sectional descriptive study was conducted based on exit surveys with AGYW held at Zimpeto and 1° de Junho Health Facilities in Maputo City, between May 1, and June 9, 2023. Data were analyzed through descriptive statistics, t-test and ANOVA, using SPSS version 20.

**Results:** 590 AGYW, aged 15-24 years of age, were included in the study. In general, knowledge of SRH and HIV services was fairly high, with knowledge of each specific service offered ranging between 38% and 97%. Knowledge about SRH and HIV services differed depending on the health facility where the AGYW sought SRH and HIV services; the participant’s age; their occupation; their religion, and who they lived with. Counseling services were the most commonly reported services attended, with >90% of participants reporting having received counseling for each of the following: STI and HIV and pregnancy prevention, sexuality, and safer sex practices. The quality of SRH and HIV services, and attitudes of the providers were considered good by >90% of AGYW. Roughly 95% of AGYW at Zimpeto Health Facility were either “satisfied” or “very satisfied”. Whereas at 1° de Junho Health Facility, only roughly 75% of AGYW were either “satisfied” or “very satisfied”, and roughly 20% of AGYW were “little satisfied that their needs had been met that day.

**Conclusions:** Among AGYW there is high levels of knowledge about counseling services in contrast to diagnostics, treatment and clinical care. Specific attention should be given to ensuring appropriate physical infrastructure, such as dedicated adolescent friendly spaces and comfortable seating, and targeted interventions designed and implemented for those health facilities’s identified. Targeted interventions should be designed and implemented for those HF’s identified with lower AGYW perceived quality of service delivery.

## Introduction

Adolescent girls and young women (AGYW) remain at the highest risk of acquiring human immunodeficiency virus (HIV) in sub-Saharan Africa (1). Globally in 2023, it was estimated that an average of 4,000 AGYW aged 15–24 years became newly infected with HIV each week, of which roughly 75% occurred in sub-Saharan Africa. AGYW are three times as likely to acquire HIV than their male counterparts (2).

Evidence shows that in sub-Saharan Africa, adolescents engage in sexual activity at a very young age (average age of 13 years), yet most do not use any form of protection against unintended pregnancies and STIs (3). In a multi-country analysis, the prevalence of first pregnancy among AGYW ranged from 7.2% (Rwanda) to 44.3% (in Congo) (4). General knowledge about sexually transmitted infections (STI) and HIV, as well as about sexual reproductive health (SRH) services has been shown to be limited among AGYW (5). Further, AGYW face a number of challenges in terms of access to comprehensive health care, meeting their contraceptive needs, and the ability to negotiate safe sex (6).

Despite various efforts by the Mozambican Ministry of Health (MoH) to promote interventions that prevent HIV infections, STI, and early pregnancy through adolescent and youth-friendly health services (AYFHS), Mozambique has the highest reported proportion of AGYW who have had sex before the age of 15 in sub-Saharan Africa. Furthermore, as of 2017, Mozambique ranks third in terms of countries with the highest birth rate among adolescents (123 per 1000 women) (7). In 2023, 61% of females in Mozambique reported having had a live birth by the age of 19 years. Mozambique is also a country with a high HIV disease burden. In 2023, the HIV prevalence ranged from 8 to 21% across Mozambique’s 11 provinces. In the same year, Maputo, the country’s capital city, reported an HIV prevalence of 16%. Nationwide, HIV prevalence among persons aged 15-24 stands at 5.4%. Within this age group, women have a higher HIV prevalence (8%) as compared to men (2.6%). While 54% of AGYW have been reported as having tested for HIV, only 1.4% received their results, which could give rise to gaps in the provision of HIV services (8).

An analysis of the gaps and obstacles in priority interventions for the prevention and treatment of HIV/AIDS in adolescents in Mozambique, carried out in 2017, found that <40% of adolescents reported having accessed AYFHS, yet only 13.6% reported actively using a modern contraceptive method (9). The most recent Demographic Health Survey (2023) carried out in Mozambique, indicates that the prevalence of modern contraceptive use was approximately 16% among AGYW (10).

Studies on the barriers to accessing SRH and HIV services across sub-Saharan Africa, commonly report inadequate information on the part of the AGYW about the availability of services, as well as their misperceptions about SRH and HIV services. In addition, services offered in an unsupportive environment and poor provider attitudes have been listed as potential barriers to accessing care (11,12).

To contribute to addressing gaps in the understanding of AGYW utilization of SRH and HIV services in Mozambique, this study aims to first, assess AGYW knowledge about SRH and HIV services availability, and second, to explore AGYW experiences with utilizing SRH and HIV services at two health facilities (HF) in Maputo City, Mozambique.

## Materials and methods

### Study design

We conducted a cross-sectional descriptive study that utilized a mixed-method approach to assess AGYW knowledge about, and experiences with, accessing SRH and HIV services in Mozambique. This analysis represents baseline (phase one) data collection of a larger multi-phase implementation science study aiming to assess the feasibility and effectiveness of an “adolescent-friendly approach” for improving access to and use of SRH and HIV services by AGYW at selected HF. At baseline, we conducted an exit survey with AGYW who had sought SRH and HIV services at the Zimpeto and 1° de Junho HFs in Maputo City, Mozambique. The selection of these health facilities was based on their prior poor performance indicators related to AGYW’s access to and use of the SRH and HIV services.

### Study population

All AGYW, aged 15-24 years, who had accessed SRH and HIV services at either of the two study facilities between May 1 and June 9, 2023, were considered eligible to participate in the exit survey. AGYW were selected by convenience and approached for enrollment as they were exiting the HF, provided they had any contact with the SRH or HIV services on the day of the interview. Initially, a sample size of 520 AGYW was defined, but during the data collection process, 598 were consented and enrolled.

### Data collection and management

Interviews took place on weekdays, during the normal service hours of the two HF (7:30 a.m. to 3:30 p.m.). The first AGYW to leave the HF during this time interval was approached and, if accepted, she was interviewed. Only after the interview was finished would other participants be approached for recruitment, and so on until the defined sample was reached. Trained interviewers, fluent in Portuguese and the principal local language, Changana, conducted the interviews using a semi-structured interview guide in a private location.

AGYW were questioned about their sociodemographic data; knowledge of the types of SRH and HIV services offered at the HF for AGYW; types of SRH and HIV services received on the day of the visit; the perceived quality of the SRH and HIV services offered on the day of the visit; and their level of satisfaction with the SRH and HIV services received on the day of the visit. Questions designed to assess AGYW knowledge about the services offered and received, were based off a list of SRH and HIV services outlined in the *Guidelines for the Implementation of adolescent and youth-friendly health services of the Mozambican Ministry of Health* (13). Questions related to the perceived quality of the services offered were based on World Health Organization (WHO) standard quality assessment questions for AYFHS (14).

A survey tool was developed using REDCap (Research Electronic Data Capture) and administered via a tablet in an offline system (REDCap v10.6.12, 2023). At the end of each day, data was synchronized and sent to a REDCap server housed at the Faculty of Medicine of University Eduardo Mondlane (UEM) in Maputo. The data were extracted from REDCap into excel format, and then imported into the Statistical Package for the Social Sciences (SPSS) Version 2.0. Data were cleaned and all lines with missing data for reference variables for analysis were excluded from the analysis.

### Data analysis

Descriptive statistics using absolute and relative frequencies were applied to identify which SRH and HIV services were known to AGYW; which SRH and HIV services were used, including modern contraceptive methods; and to assess AGYW perceptions of the environment of the health facility to serve adolescents. T-test and ANOVA were applied to identify the sociodemographic factors associated with average knowledge, considering a significance level of 5%. To assess the average knowledge of participants, each of the 10 questions relating to each type of SRH and HIV service was assigned a score of 10 percent. Afterwards, the knowledge level variable was created, which adds up the score for the number of services known by each participant, ranging from 0 to 100%. The average knowledge was calculated from the knowledge level variable. The average knowledge was compared between the AGYW in the two study sites and their sociodemographic characteristics to identify the factors associated with the level of average knowledge.

### Ethical considerations

The study protocol was reviewed and approved by the Mozambican National Bioethics Committee for Health (Ref: 88/CNBS/23). Administrative approval was granted by the MoH of Mozambique (Note nr: 396/GMS/290/023). All participants provided written informed consent prior to participation in the exit interview. For participants below the age of 18 years, informed consent was first obtained from a parent/guardian and then informed assent from the participant. No identifying information was recorded by the interviewer to ensure anonymity.

## Results

### Participant characteristics

A total of 598 AGYW were approached and surveyed. Eight participants were subsequently excluded from the analysis because of missing data. A total of 590 AGYW were included in this analysis, of which 304 (51.5%) were recruited from the 1° de Junho HF and 286 (48.5%) from the Zimpeto HF. Age of the AGYW’s was nearly evenly split between those aged 15-19 years (51%) and those 20-24 years (49%), with a mean age of 20 years ±2.42 for all participants. Approximately 70% (n=419) of respondents listed themselves as a current student, of which the majority (86.8%) had achieved a level of secondary education. Of the AGYW interviewed, 79.6% (n=468) were single, while 20.4% (n=122) were married or in a non-civil marriage. Christianity was the leading religion, being the majority classified as Christian-Other (which included protestant and evangelical). The majority of participants (65.5%) lived with their parents, while 19.3% (n=114) lived with their husbands (see Table 1).

**Table 1:** Participant Sociodemographic Characteristics.

| Characteristic (n=590) | n (%) |
| --- | --- |
| Health facility |  |
| 1° de Junho HF | 304 (51.5) |
| Zimpeto HF | 286 (48.5) |
| Age |  |
| 15-19 years | 301 (51.0) |
| 20-24 years | 289 (49.0) |
| Mean age | 20±2.42 |
| Level of education |  |
| No education | 27 (4.6) |
| Primary | 31 (5.3) |
| Secondary | 513 (86.8) |
| Higher | 19 (3.2) |
| Current student |  |
| Yes | 414 (70.2) |
| Marital status |  |
| Married/Marital Union | 122 (20.4) |
| Single | 468 (79.6) |
| Religion |  |
| Christian - Catholic | 203 (34.4) |
| Christian - other | 316 (53.5) |
| Muslim | 29 (4.9) |
| No religion or other | 42 (7.1) |
| Who you live with? |  |
| Alone | 6 (1.0) |
| Mother/Father/Parents | 386 (65.4) |
| Siblings | 19 (3.2) |
| Grandfather/Grandmother | 30 (5.1) |
| Uncles | 35 (5.9) |
| Husband | 114 (19.3) |

### AGYW knowledge of sexual reproductive health and HIV services available at health facility

In general, AGYW knowledge of SRH and HIV services was fairly high, with knowledge of each specific service offered ranging between 38% and 97%. The best-known SRH and HIV services were pregnancy prevention counseling (97%); HIV and STI prevention counseling (97%); safe sex counseling (97%); sexuality counseling (96%); HIV and STI diagnostic testing (89%); antenatal care consultations (84%); and gender-based violence (GBV) services (82%). Pregnancy testing services (64%), and abortion services (72%) were moderately known, and the least known service was the postpartum care clinics (38%) (see Fig 1).

Average knowledge about SRH and HIV services differed depending on where the AGYW sought healthcare, the participants age, their status of status of employment, their religion, and who they lived with. AGYW who attend the Zimpeto HF had a higher average level of knowledge about the SRH and HIV services offered (86.6%; CI: 83.8-89.3) compared to those who attended the 1° de Junho HF (76.3%; CI:74.3-78.3) (p<0.001). AGYW aged 20-24 years had a higher average level of knowledge (83%; CI: 80.7-85.4), when compared to AGYW aged 15-19 years (79.6%; CI: 77.1-82.1) (p=0.046). AGYW who are actively in school had a higher average level of knowledge about SRH and HIV services (82.4%; CI: 80.2-84.5), compared to adolescents who are out of school (78.7%; CI: 75.8-81.7) (p=0.048). AGYW who report a religious affiliation: Protestant (77.9%; CI: 75.5-80.3), Catholic (86.5%; CI:83.8-89.3), and Muslim (88.9%; CI: 83.2-94.7), had a higher average level of knowledge when compared to those reporting no religious affiliation (75.8%; CI: 67.9-83.8) (p<0.001). Of note, a significantly higher average level of knowledge was observed among Muslim and Catholic participants, compared to Protestants or non-religious counterparts. Lastly, AGYW who live with their parents had a higher average level of knowledge (84.20% CI: 82.2-86.2) when compared to other groups (living alone, with siblings, grandmother/grandfather, uncles or with spouses, respectively), (78.3% CI: 66.1 – 90.6; 81.6% CI: 74.2 – 89.0; 69.3% CI: 57.8 – 80.9; 69.7% CI: 60.9 – 78.5 and 78.3% CI: 74.5 – 81.9) (p<0.001). All associations observed were statistically significant. The characteristics not showing a statistically significant association with knowledge about SRH and HIV services were marital status and level of education (see Table 2).

**Table 2:** Factors associated with knowledge about SRH and HIV services offered.

| Characteristics | n | Mean | 95% CI | *p-value* |
| --- | --- | --- | --- | --- |
| Site |  |  |  | <0.001* |
| 1° de Junho HF | 304 | 73.3 | 74.3 – 78.3 |  |
| Zimpeto HF | 286 | 86.6 | 83.8 – 89.3 |  |
| Age |  |  |  | 0.046* |
| 15-19 years | 301 | 79.6 | 77.1 – 82.1 |  |
| 20-24 years | 289 | 83.1 | 80.7 – 85.6 |  |
| Level of education |  |  |  | 0.089 |
| No schooling | 27 | 83.7 | 72.7 - 94.7 |  |
| Primary | 31 | 72.9 | 63.9 - 81.9 |  |
| Secondary | 513 | 81.9 | 80.0 - 83.7 |  |
| Higher | 19 | 76.3 | 70.5 - 82.2 |  |
| Employment Status<sup>#</sup> |  |  |  | 0.048* |
| Student | 414 | 82.4 | 80.3 - 84.5 |  |
| Unemployed/Out of School | 176 | 78.7 | 75.7 – 81.7 |  |
| Marital status |  |  |  | 0.118 |
| Single | 468 | 81.8 | 79.9 - 83.8 |  |
| Married/marital union | 117 | 78.4 | 74.8 – 82.0 |  |
| Religion |  |  |  | <0.001* |
| No religion | 41 | 75.9 | 67.9 – 83.8 |  |
| Protestant | 316 | 77.9 | 75.6 – 80.3 |  |
| Catholic | 203 | 86.6 | 83.8 – 89.3 |  |
| Muslim | 29 | 89.0 | 83.2 – 94.8 |  |
| Who do you live with? |  |  |  | <0.001* |
| Alone | 6 | 78.3 | 66.1 – 90.6 |  |
| Mother/Father | 386 | 84.2 | 82.2 – 86.2 |  |
| Siblings | 19 | 81.6 | 74.2 – 89.0 |  |
| Grandmother/Grandfather | 30 | 69.3 | 57.8 – 80.9 |  |
| Uncles | 35 | 69.7 | 60.9 – 78.5 |  |
| Spouse | 114 | 78.3 | 74.5 – 81.9 |  |

### Types of SRH and HIV services received by AGYW on the day of the health facility visit

AGYW reported a variety of SRH and HIV services they sought on the day of their interview. Counseling services were the most commonly reported services, with >90% of participants reporting having received counseling for each of the following: STI and HIV prevention, pregnancy prevention, sexuality, and safer sex practices. Further, 51% of participants attended Family Planning services and 31% received testing for HIV and/or another STI. Smaller numbers of participants attended other services such as antenatal care or gender-based violence (GBV) counseling (<20% for each service). Among those participants who attended Family Planning services on the day of visit to the HF (n=303), birth control options were received in the following proportions: 33% injectables (Depo-Provera); 30% oral contraceptive pill; 16% male condoms; 12% female condoms; and 10% implant. Participants could receive more than one type of contraceptive (see Fig 2).

### AGYW perceptions of the quality of SRH and HIV services offered at their health facility

AGYW perceptions based on their experiences accessing SRH and HIV services were captured through a series of questions to explore their perceptions of the quality of care received, including their opinion about the physical environment in which the services were offered. Overall, more than 90% of participants reported to have received counseling in private spaces and that upon arrival to the service, they were greeted and served according to their needs. At Zimpeto HF, over 95% of participants reported that services were offered in a dedicated space catering to adolescents. In contrast, roughly 15% of participants at 1° de Junho HF reported a lack of an adolescent dedicated space. In addition, the vast majority of participants (92.6%) at 1° de Junho HF reported a lack of separate waiting room for adolescents, compared to Zimpeto HF, where <10% of participants reported a lack a separate waiting room. At both facilities, >90% of participants reported that the HF did not have a dedicated schedule for when adolescent services were available and >25% of AGYW reported no comfortable sitting arrangements at the waiting area.

In terms of service quality, >90% of participants at both facilities ranked the attitude of the provider as “Good”. At Zimpeto HF >90% of participants ranked the quality of services provided as “Good”, whereas at 1° de Junho HF, only 79% ranked the services as “Good” and 18% ranked them only as acceptable.

When asked about how satisfied the participant was with regards to if their needs had been met that day, roughly 95% of AGYW at Zimpeto HF responded they were either “satisfied” or “very satisfied”. Whereas at 1° de Junho HF, roughly 75% of AGYW reported they were either “satisfied” or “very satisfied”, and approximately 20% responded they were “little satisfied” (see Table 3).

**Table 3:** AGYWs’ experiences and perceptions about SRH and HIV services offered.

| How do you evaluate the environment of the health facility’s services to adolescents? | 1° de Junho<br>n (%) | Zimpeto<br>n (%) | Total<br>n (%) |
| --- | --- | --- | --- |
| Is there a counseling area for adolescents that provides privacy? |  |  |  |
| No | 23 (7.6) | 6 (2.1) | 29 (4.9) |
| Yes | 281 (92.4) | 280 (97.9) | 561 (95.1) |
| Are adolescents greeted and served according to their needs or those of their partner(s)? |  |  |  |
| No | 27 (8.9) | 8 (2.8) | 35 (5.9) |
| Yes | 277 (91.1) | 278 (97.2) | 555 (94.1) |
| Does the health facility have a separate space to offer services to adolescents? |  |  |  |
| No | 48 (15.8) | 14 (4.9) | 62 (11.0) |
| Yes | 256 (84.2) | 272 (95.1) | 528 (89.0) |
| Does the health facility have a comfortable place for adolescents to sit? |  |  |  |
| No | 109 (35.9) | 70 (24.5) | 178 (30.3) |
| Yes | 195 (64.1) | 216 (75.5) | 411 (69.7) |
| Does the health facility have a separate waiting room for adolescents? |  |  |  |
| No | 276 (92.6) | 22 (7.4) | 298 (51.0) |
| Yes | 28 (9.6) | 264 (90.4) | 292 (49.0) |
| Is there a specific schedule for adolescents in this health facility? |  |  |  |
| No | 281 (92.5) | 285 (99.7) | 566 (95.9) |
| Yes | 23 (7.5) | 1(0.03) | 24 (4.1) |
| How do you evaluate the attitude of the provider who assisted you? |  |  |  |
| Bad | 10 (3.3) | 0 (0.0) | 10 (1.7) |
| Acceptable | 40 (13.2) | 9 (3.1) | 49 (8.3) |
| Good | 254 (83.6) | 277 (96.9) | 531 (90.0) |
| Overall, how do you evaluate the provision of SRH and HIV services in this health facility? |  |  |  |
| Bad | 9 (3.0) | 3 (1.0) | 12 (2.0) |
| Acceptable | 55 (18.1) | 25 (8.7) | 80 (13.6) |
| Good | 240 (79.0) | 258 (90.2) | 498 (84.4) |
| How satisfied were you that your needs were met today? |  |  |  |
| Little satisfied | 62 (20.4) | 12 (4.2) | 74 (12.5) |
| Not satisfied | 17 (5.6) | 3 (1.2) | 20 (3.4) |
| Satisfied | 193 (63.5) | 258 (90.0) | 451 (76.4) |
| Very satisfied | 32 (10.5) | 13 (4.6) | 42 (7.7) |

Note: The NO option includes those who answered that they don't know

## Discussion

This study aimed to assess the knowledge of AGYW seeking health services in selected HF in Maputo, Mozambique related to the SRH and HIV services being offered, as well as to explore their experiences in utilizing these services. This type of assessment is relevant and addresses the WHO recommendation to ensure that adolescents are aware of what health services are being provided, and where, when, and how to obtain them (14).

This study represents a baseline assessment of a larger implementation science study to evaluate the feasibility and effectiveness of an “adolescent-friendly approach” for improving access to and use of SRH and HIV services by AGYW.

Broadly speaking, knowledge about SRH and HIV services in our population of Mozambican AGYW was high. Specifically, the best-known services were counseling services at the HFs related to safe sexual practices including pregnancy prevention, HIV/STI prevention, and understanding one’s sexuality. Awareness of testing services for pregnancy and HIV/STI was slightly less, and the least known services were those related to antenatal and post-partum care. When we compare our AGYW population to other similar populations across sub-Saharan Africa, our findings show that generalized knowledge about SRH and HIV services in Mozambican AGYW is higher (15–18). However, when we begin looking at the different types of services individually, we found that knowledge of HIV and STI testing was generally higher in Mozambique as well as in other countries such as Ghana, Ethiopia, and Nigeria. The high level of knowledge observed in the group involved in our study, compared to the groups involved in other studies carried out in Ghana, Ethiopia, and Nigeria, may be linked to the fact that our study was based in the HF and the group involved had just had contact with SRH and HIV services. Whereas the studies carried out in other contexts were based at community level, possibly with a memory bias, which can limit the level of knowledge of SRH and HIV services. Furthermore, relatively low Mozambican AGYW knowledge about GBV services offered is consistent with a similar low knowledge seen in these other countries (15,19,20).

We found a significant association between knowledge and increased age, being a student, religion, the HF where services were sought, and whom one lives with. These findings are not surprising. First, with increased age, AGYW gain the autonomy to decide about their health and likely have had more opportunities to visit a HF and more life experiences that would enhance their overall knowledge about these services. Second, AGYW students are more likely to have more knowledge compared to those who don’t, since school is one of the major sources of health information (19–22). There is controversial evidence about the association between religious affiliation and the level of knowledge about sexual and reproductive health services and HIV. While other studies suggest that religious affiliation is a protective factor against the risk of HIV (23), there is recognition that religion may offer inadequate information related to SRH and HIV (24), and at times has a negative influence on the level of knowledge about SRH among adolescents (25). Our results show a significant difference in the level of knowledge of SRH and HIV between AGYW who attend the Zimpeto HF and those who attend the 1° de Junho HF, which suggests the need to delve deeper into the internal factors of each center to better explain these differences. Furthermore, associations between the level of knowledge of SRH and HIV and the health center where AGYW seek services, may be linked to the fact that this study was carried out at the HF and involved only AGYW who visited the HF and had just had contact with SRH and HIV Services.

Exposure to information about SRH services has been reported as one of the predictors of SRH utilization (15,19,26). However, in this study, this seemed to be the opposite. Among the AGYW who visited health centers and had access to SRH counseling services, few of them had access to STI, HIV and early pregnancy prevention supplies. Similar results related to low use of HIV testing services; STI treatment; family planning among AGYW was found in another studies conducted in similar context (7,15,20,21,27,28).

Study participants were generally favorable about the quality of the SRH and HIV services they had accessed in terms of existence of separate and visible areas for AGYW, and the attitude of the providers they had seen. Similar results on quality of the SRH and provider attitude were found in a study evaluating the quality of AYFHS carried out in Mozambique, Ethiopia and Nigeria (29–31), but different from the results of studies carried out in Mexico (32) and Nigeria (33), who reported negative provider attitudes characterized by judgment of AGYW when they seek SRH and HIV services. However, our results suggest that the conditions of the waiting rooms at the HFs was not comfortable. Further, there was evidence of the lack of a specific schedule for when adolescent services are offered. Nevertheless, the guide to implementing a standards approach to improving the quality of health services for adolescents, recommends that adequate seating should be made available in the waiting room for the normal flow of patients. It also recommends that HFs should have convenient hours of operation that facilitates adolescents access to these health services (14).

Despite these complaints, the high level of reported satisfaction among the participants seems to indicate that the quality of the service provided for AGYW is perceived as good. A similar high level of satisfaction was found in assessments of AYFHS from both Eastern and Southern Africa Regions (7,29,34,35).

## Conclusion

Knowledge about SRH and HIV services offered in HFs is inconsistent across the different types of services, with high levels of knowledge about counseling in contrast to diagnostics, treatment and clinical care. The results of this study suggest the need to balance the dissemination of the different SRH and HIV services targeting AGYW in the catchment areas involved in the study. Additionally, site specific attention should be given to ensuring appropriate physical infrastructure exists that takes into account the unique needs of AGYW, such as dedicated adolescent friendly spaces and comfortable seating. Finally, targeted interventions should be designed and implemented for those HF’s identified with lower AGYW perceived quality of service delivery, such as we are proposing for the 1° de Junho HF.

## Data Availability

The data underlying the results presented in the study are available from

## Autho’s contributions

VM designed the study, guided by KM, BC and TDM. VM collected the data. VM, LH, and KTK analyzed the data. VM drafted the manuscript. All authors contributed to interpretation of results and revising the manuscript.

## Acknowledgements

The authors would like to thank the Department of Community Health, Faculty of Medicine, Eduardo Mondlane University; Department of Tropical Medicine and Infectious Diseases, School of Public Health and Tropical Medicine, Tulane University. The Health Managers of 1° de Junho and Zimpeto Health Facilities and the leadership of Adolescents and Youth Friendly Service at both health facilities.

---

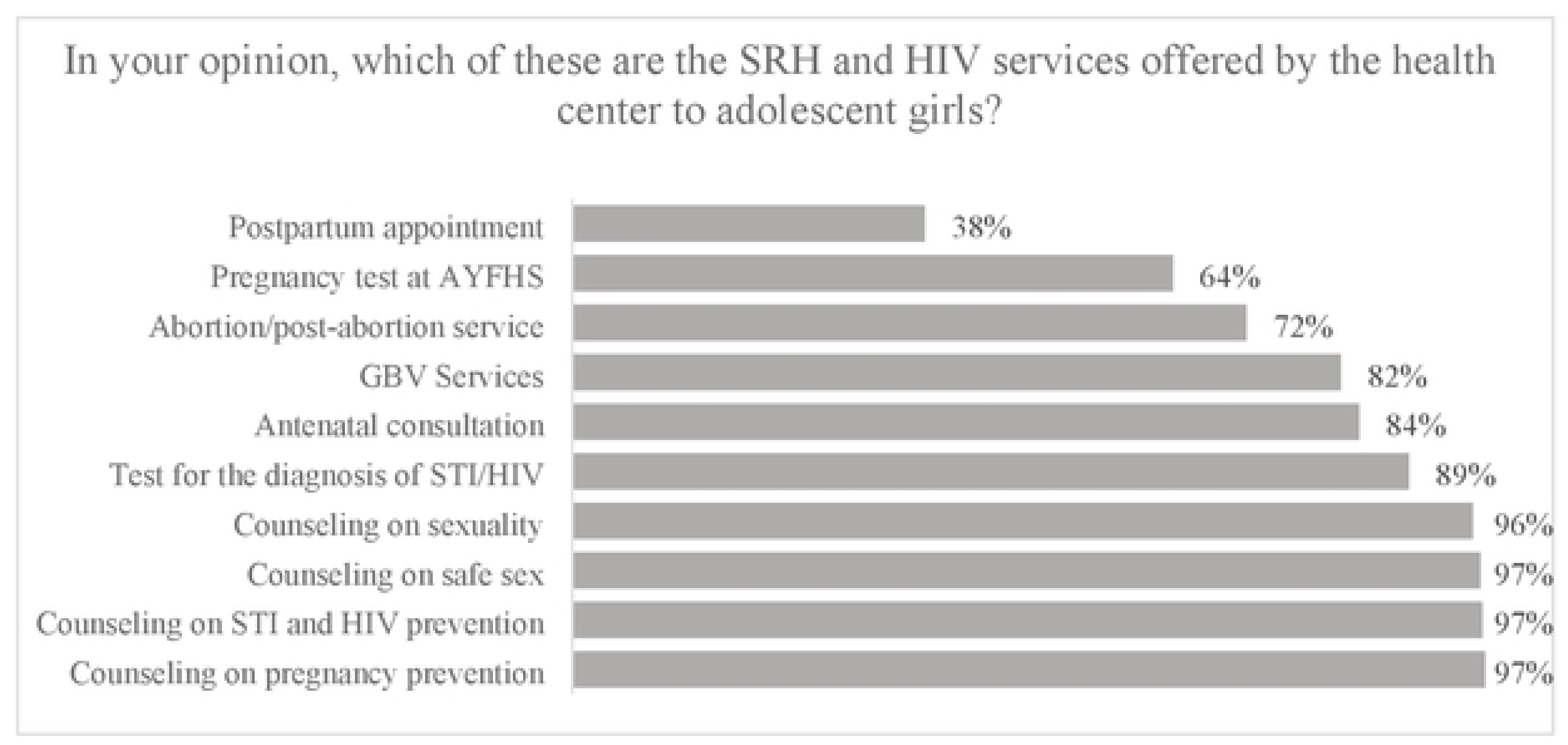

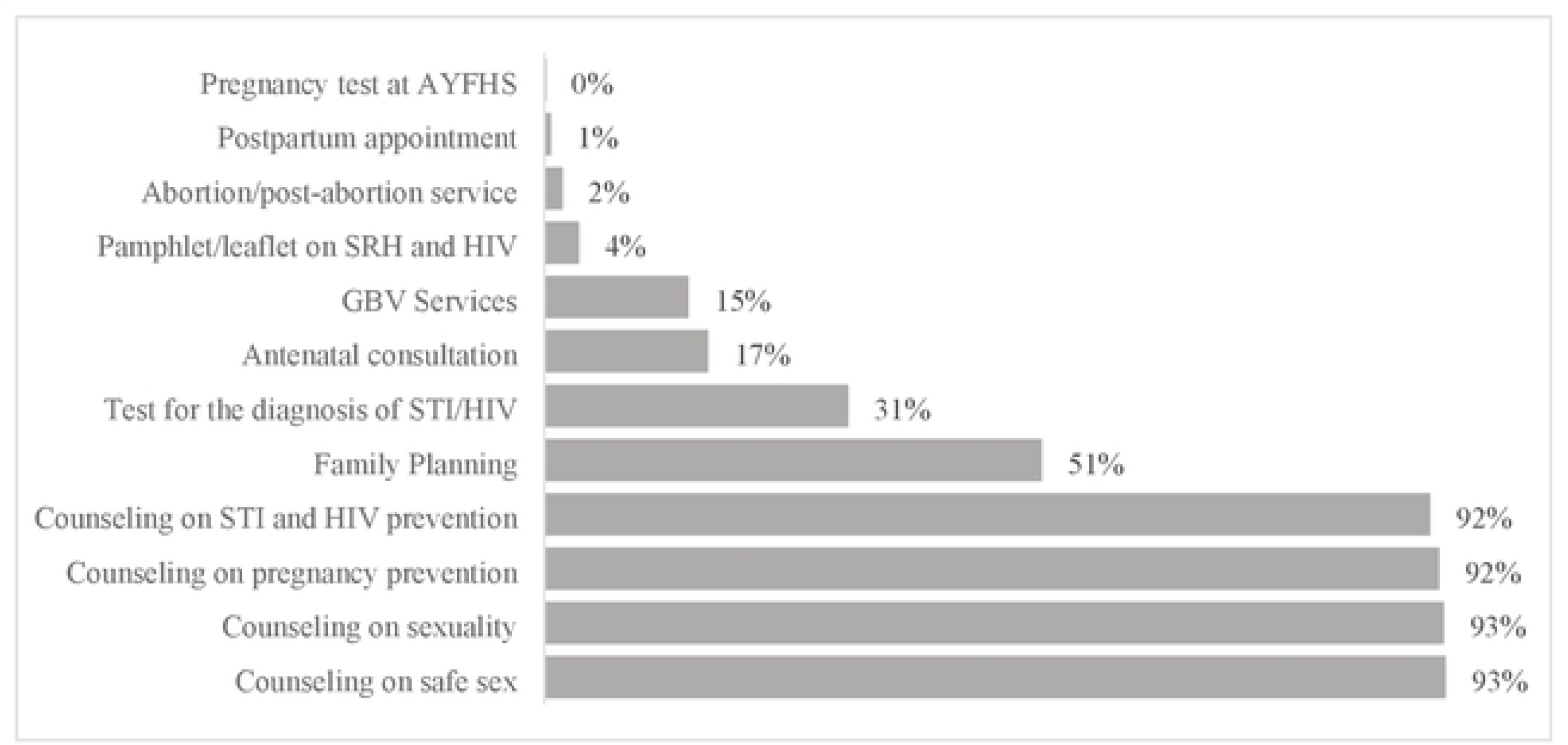

## Bibliography

1. Murewanhema G, Musuka G, Moyo P, Moyo E, Dzinamarira T. HIV and adolescent girls and young women in sub-Saharan Africa: A call for expedited action to reduce new infections. IJID Reg [Internet]. 2022;5(August):30–2. Available from: 10.1016/j.ijregi.2022.08.009

2. UNAIDS. REGIONAL PROFILE; EASTERN AND SOUTHERN AFRICA [Internet]. Vol. 2024. 2024. Available from: https://www.unaids.org/en/resources/fact-sheet

3. James PB, Osborne A, Babawo LS, Bah AJ, Margao EK. The use of condoms and other birth control methods among sexually active school-going adolescents in nine sub-Saharan African countries. BMC Public Health. 2022 Dec 1;22(1).

4. Ahinkorah BO, Kang M, Perry L, Brooks F, Hayen A. Prevalence of first adolescent pregnancy and its associated factors in sub-Saharan Africa: A multi-country analysis. PLoS One [Internet]. 2021;16(2 February):1–16. Available from: 10.1371/journal.pone.0246308

5. Finlay JE, Assefa N, Mwanyika-Sando M, Dessie Y, Harling G, Njau T, et al. Sexual and reproductive health knowledge among adolescents in eight sites across sub-Saharan Africa. Trop Med Int Heal. 2020 Jan 1;25(1):44–53.

6. Yah CS, Ndlovu S, Kutywayo A, Naidoo N, Mahuma T, Mullick S. The prevalence of pregnancy among adolescent girls and young women across the Southern African development community economic hub: A systematic review and meta-analysis. Vol. 10, Health Promotion Perspectives. Tabriz University of Medical Sciences; 2020. p. 325–37.

7. UNFPA & IPPF. Regional Report: Assessment of Adolescents and Youth-Friendly Health Service Delivery EAST AND SOUTHERN AFRICA REGION [Internet]. Johannesburg and Nairobi; 2017. Available from: https://healtheducationresources.unesco.org/library/documents/regional-report-assessment-adolescents-and-youth-friendly-health-service-delivery

8. MISAU & INS. Inquérito Nacional Sobre o Impacto do HIV e SIDA em Moçambique [Internet]. Maputo, Moçambique; 2023. Available from: http://ins.gov.mz

9. CNCS, UNICEF, ONUSIDA. Análise aprofundada das lacunas e obstáculos das intervenções prioritárias de prevenção e tratamento do HIV / SIDA em adolescentes em Moçambique. Maputo, Moçambique; 2017.

10. INE. Inquérito Demográfico e de Saúde 2022-23 [Internet]. Vol. 23. 2023. Available from: https://www.ine.gov.mz/web/guest/d/ids-2022-23-relatorio-final

11. Sidamo NB, Kerbo AA, Gidebo KD, Wado YD. Socio-Ecological Analysis of Barriers to Access and Utilization of Adolescent Sexual and Reproductive Health Services in Sub-Saharan Africa: A Qualitative Systematic Review. Open Access J Contracept. 2023 Jun;Volume 14:103–18.

12. Zepro NB, Ali NT, Tarr N, Medhanyie AA, Paris DH, Probst-Hensch N, et al. Sexual and reproductive health services use among adolescents in pastoralist settings, northeastern Ethiopia. BMC Health Serv Res. 2023 Dec 1;23(1).

13. MISAU. Linhas de Orientação para implementação de Serviços Amigos de Adolescentes e Jovens (SAAJ). Maputo, Moçambique; 2023. p. 51.

14. UNAIDS. Global standards for quality health-care services for adolescents: a guide to implement a standards-driven approach to improve the quality of health care services for adolescents. World Heal Organ [Internet]. 2015;1:1–40. Available from: http://apps.who.int/iris/bitstream/10665/183935/1/9789241549332_vol1_eng.pdf

15. Akakpo E, Sah C, Kumah A, Fianu PL, Oppong DA, Kodjo MM. Quality Health Services for Adolescents: Assessing Awareness and Use of Adolescent Sexual Reproductive Health Services in Keta, Ghana. Glob J Qual Saf Healthc. 2024 Feb 26;

16. Ninsiima LR, Chiumia IK, Ndejjo R. Factors influencing access to and utilisation of youth-friendly sexual and reproductive health services in sub-Saharan Africa: a systematic review. Vol. 18, Reproductive health. NLM (Medline); 2021. p. 135.

17. Abiodun O, Sotunsa J, Jagun O, Faturoti B, Ani F, John I, et al. Prevention of unintended pregnancies in Nigeria; the effect of socio-demographic characteristic on the knowledge and use of emergency contraceptives among female university students. Int J Reprod Contraception, Obstet Gynecol. 2015;4(3):755–64.

18. O. Ajike S. Adolescent/Youth Utilization of Reproductive Health Services: Knowledge Still a Barrier. J Fam Med Heal Care. 2016;2(3):17.

19. Abdurahman C, Oljira L, Hailu S, Mengesha MM. Sexual and reproductive health services utilization and associated factors among adolescents attending secondary schools. Reprod Health. 2022 Dec 1;19(1).

20. Belay HG, Arage G, Degu A, Getnet B, Necho W, Dagnew E, et al. Youth-friendly sexual and reproductive health services utilization and its determinants in Ethiopia: A systematic review and meta-analysis. Heliyon. 2021 Dec 1;7(12).

21. Ugwu NH, Igwe I, Nwokeoma BN, Ajuzie HD, Iwuamadi KC, Ezike SC, et al. Adolescents’ knowledge and use of sexual and reproductive health services in the Federal Capital Territory, Nigeria. Afr J Reprod Health. 2022;26(6):80–8.

22. Eze II, Mbachu CO, Agu IC, Akamike IC, Eigbiremolen G, Onwujekwe O. Determinants of awareness, value perception, and societal support for sexual and reproductive health services among in-school adolescents in South-eastern Nigeria. BMC Health Serv Res. 2023;23(1):1–10.

23. Shaw SA, El-Bassel N. The Influence of Religion on Sexual HIV Risk. AIDS Behav. 2014;18(8):1569–94.

24. Tafesse W, Chalkley M. Faith-based provision of sexual and reproductive healthcare in Malawi. Soc Sci Med [Internet]. 2021;282(May):113997. Available from: 10.1016/j.socscimed.2021.113997

25. Gebresilassie K, Boke M, Yenit M, Baraki A. Knowledge level and associated factors about sexual and reproductive health rights among University of Gondar students, Gondar Ethiopia. Int J Sex Reprod Heal Care [Internet]. 2019 Aug 30;2(1):016–20. Available from: https://www.reprodgroup.us/articles/IJSRHC-2-106.php

26. Agbenu I, Kyei J, Naab F. Female adolescent sexual reproductive health service utilization concerns: A qualitative enquiry in the Tema metropolis of Ghana. PLoS One. 2024 Feb 1;19(2 February).

27. Getachew S, Abate L, Asres A, Mandefro A. Knowledge, Attitude, and Practice toward Youth-Friendly Reproductive Health Services among Mizan-Tepi University Students, South-Western Ethiopia. Sci World J. 2022;2022.

28. Utaka EN, Sekoni AO, Badru FA. Knowledge and utilization of sexual and reproductive health services among young males in a slum area in Nigeria: A cross-sectional study. Heliyon. 2023 Jun 1;9(6).

29. Bomfim E, Mupueleque MA, Dos Santos DMM, Abdirazak A, Bernardo R de A, Zakus D, et al. Quality assessment in primary health care: Adolescent and youth friendly service, a mozambican case study. Pan Afr Med J. 2020 Sep 1;37:1–10.

30. Tilahun T, Bekuma TT, Getachew M, Seme A. Assessment of access and utilization of adolescent and youth sexual and reproductive health services in western Ethiopia. Reprod Health. 2021 Dec 1;18(1).

31. Alamdo AG, Debelle FA, Gatheru PM, Manu A, Enos JY, Yirtaw TG. Youth-friendly health service in Ethiopia: Assessment of care friendliness and user’s satisfaction. PLoS One [Internet]. 2024;19(7 July):1–18. Available from: 10.1371/journal.pone.0307142

32. Pastrana-Sámano R, Heredia-Pi IB, Olvera-García M, Ibáñez-Cuevas M, De Castro F, Hernández AV, et al. Adolescent Friendly Services: Quality assessment with simulated users. Rev Saude Publica. 2020;54:1–11.

33. Arije O, Madan J, Hlungwani T. Quality of sexual and reproductive health services for adolescents and young people in public health facilities in Southwest Nigeria: a mystery client study. Glob Health Action. 2022;15(1).

34. Godia PM, Olenja JM, Hofman JJ, Van Den Broek N. Young people’s perception of sexual and reproductive health services in Kenya. BMC Health Serv Res. 2014 Apr 15;14(1).

35. Mulugeta B, Girma M, Kejela G, Meskel FG, Andarge E, Zerihun E. Assessment of Youth-Friendly Service Quality and Associated Factors at Public Health Facilities in Southern Ethiopia: A Facility-Based Cross-Sectional Study. Biomed Res Int. 2019;2019.

